# Automating postmarketing pharmacovigilance with large language model agents

**DOI:** 10.64898/2026.11.10.26367711

**Authors:** Ruiheng Zhang, Yekai Zhou, Ian CK Wong, Celine SL Chui, Doris Yu, Zhenqin Wu, Ruibang Luo

**Author notes:** **Corresponding Authors:** Dr. Yekai Zhou, 5/F Academic Building, 3 Sassoon Road, Pokfulam, Hong Kong Special Administration Region, China., Prof. Ruibang Luo, Rm 301 Chow Yei Ching Building, The University of Hong Kong, Pokfulam Road, Pokfulam, Hong Kong Special Administration Region, China. **Co-first authors** (Ruiheng Zhang, Yekai Zhou).

## Abstract

Postmarketing pharmacovigilance relies on spontaneous adverse event reporting and labor-intensive observational studies, which suffer from underreporting and delayed risk detection. Large language model (LLM) agents could automate real-world evidence generation, but generalist agents make ad hoc analytical decisions that compromise epidemiological validity and reproducibility. Here we present MedCheckpoint, a multi-agent LLM system for automated postmarketing pharmacovigilance. MedCheckpoint grounds each analytical stage in a curated, source-traceable medical knowledge base, locks core study-design parameters before execution, performs patient-level analyses with human-verified deterministic statistical tools, and enforces checkpoint rules that abort infeasible analyses, with optional human-in-the-loop review. We evaluated MedCheckpoint using a 54-question benchmark on a dual-purpose simulated electronic health record (EHR) testbed of 2.7 million individuals, designed with reliable clinical signals to evaluate routine analytical accuracy alongside spurious signals to stress-test system rigor. Across the benchmark, MedCheckpoint outperformed two coding-agent baselines in report quality across five dimensions rated by three independent LLM judges (overall means 4.81–4.88 versus 4.25–4.55 and 4.02–4.29) and yielded higher run-to-run consistency (mean within-question standard deviation 0.116– 0.186 versus up to 0.440). Crucially, while baselines failed on up to 2 of 21 runs when handling infeasible comparisons or unmeasurable outcomes, MedCheckpoint produced no misleading failures across all seven stress-test trap questions with known data limitations. MedCheckpoint enables reliable, auditable, and scalable pharmacovigilance analysis.

## Introduction

Randomized controlled trials are the gold standard for establishing drug efficacy, but their limited sample sizes, short follow-up, and restrictive eligibility criteria leave rare, delayed, and subgroup-specific adverse effects undetected^1,2^. Postmarketing pharmacovigilance is therefore essential for identifying risks that emerge only in routine clinical use^1^. Current practice relies mainly on spontaneous adverse event reporting and manually conducted observational studies^2,3^. Spontaneous reporting suffers from severe underreporting and reporting bias^2,4^, whereas observational studies are typically reactive and hypothesis-driven, requiring custom cohort construction, statistical analysis, and expert interpretation for each safety question^3,5^. This case-by-case paradigm consumes substantial time and specialized expertise, fundamentally limiting the scale and timeliness of drug safety surveillance, particularly for long-term treatment effects that may take years to manifest^1,3,6^.

LLM agents offer a route to break this scalability bottleneck by automating complex analytical workflows. In related clinical domains, LLM agents have been shown to perform clinical calculations with curated toolkits^7^, to assist the design of randomized controlled trials^8^, and to complete multi-step clinical tasks in controlled evaluations^9^. However, automating postmarketing pharmacovigilance poses a distinct methodological challenge. Proactive surveillance tests exposure to one drug against many clinical outcomes, producing long multi-task workflows that rapidly exhaust the LLM’s input context^10^. Under this load, generalist agents improvise analytical decisions mid-workflow^11^, and prompt-based instructions alone cannot enforce epidemiological rules across extended execution chains. Such improvisation compounds into silent protocol departures, omitted statistical tests, and discrepancies between planned and executed analyses—failure modes that are unacceptable in clinical and regulatory settings, where an irreproducible decision can directly alter a safety conclusion.

Here we present MedCheckpoint, a multi-agent LLM system designed to automate postmarketing pharmacovigilance under explicit epidemiological constraints. MedCheckpoint organizes a curated medical knowledge base by analytic stage—query interpretation, cohort specification, analysis execution, and report generation—so that each agent decision is grounded in source-traceable, human-verified evidence rather than improvised. A protocol lock prospectively fixes core study-design parameters before analysis begins. Patient-level computations are delegated to human-verified deterministic statistical tools instead of code generated at runtime, and a checkpoint engine records execution status and aborts analyses that fail predefined feasibility rules, with optional human-in-the-loop review at critical decision points. In this architecture, the LLM serves as an interface and coordinator, while the decisions that determine validity are governed by verified knowledge and prespecified rules.

We evaluated MedCheckpoint on a benchmark of 54 natural-language drug safety questions using a dual-purpose simulated electronic health record (EHR) from 2.7 million individuals, which is designed with reliable clinical signals to evaluate routine analytical accuracy alongside spurious signals to stress-test system rigor. MedCheckpoint is evaluated against two baselines sharing the same backbone LLM: a general-purpose coding agent, and the same agent supplied with detailed methodological guidance. We used LLM judges to support scalable, automated assessment of report quality. Across five quality dimensions rated by three independent LLM judges, MedCheckpoint achieved the highest report quality and the most consistent outputs across repeated runs. On seven questions with known data limitations, it produced no misleading failure, correctly aborting infeasible comparisons while baselines occasionally substituted general medical knowledge for data analysis. A case study on effect modification by pre-existing diabetes illustrates how the protocol lock and deterministic tools prevent the mid-analysis revisions that compromised baseline analyses. These results demonstrate that constraining LLM agents with verified knowledge and deterministic workflows delivers reliable and auditable pharmacovigilance analysis at scale.

## Results

### Overview of MedCheckpoint workflow

MedCheckpoint structures exploratory EHR-based pharmacovigilance into three stages: protocol generation by the Case Generator, analysis coordinated by the Analyst, and reporting by the Report Writer (Fig. 1a). In the first stage, the system converts a natural language drug safety question into a structured study protocol. The protocol defines the drug exposure, comparator when applicable, target population, safety outcomes, covariates, and statistical analysis plan.

**Fig. 1.**
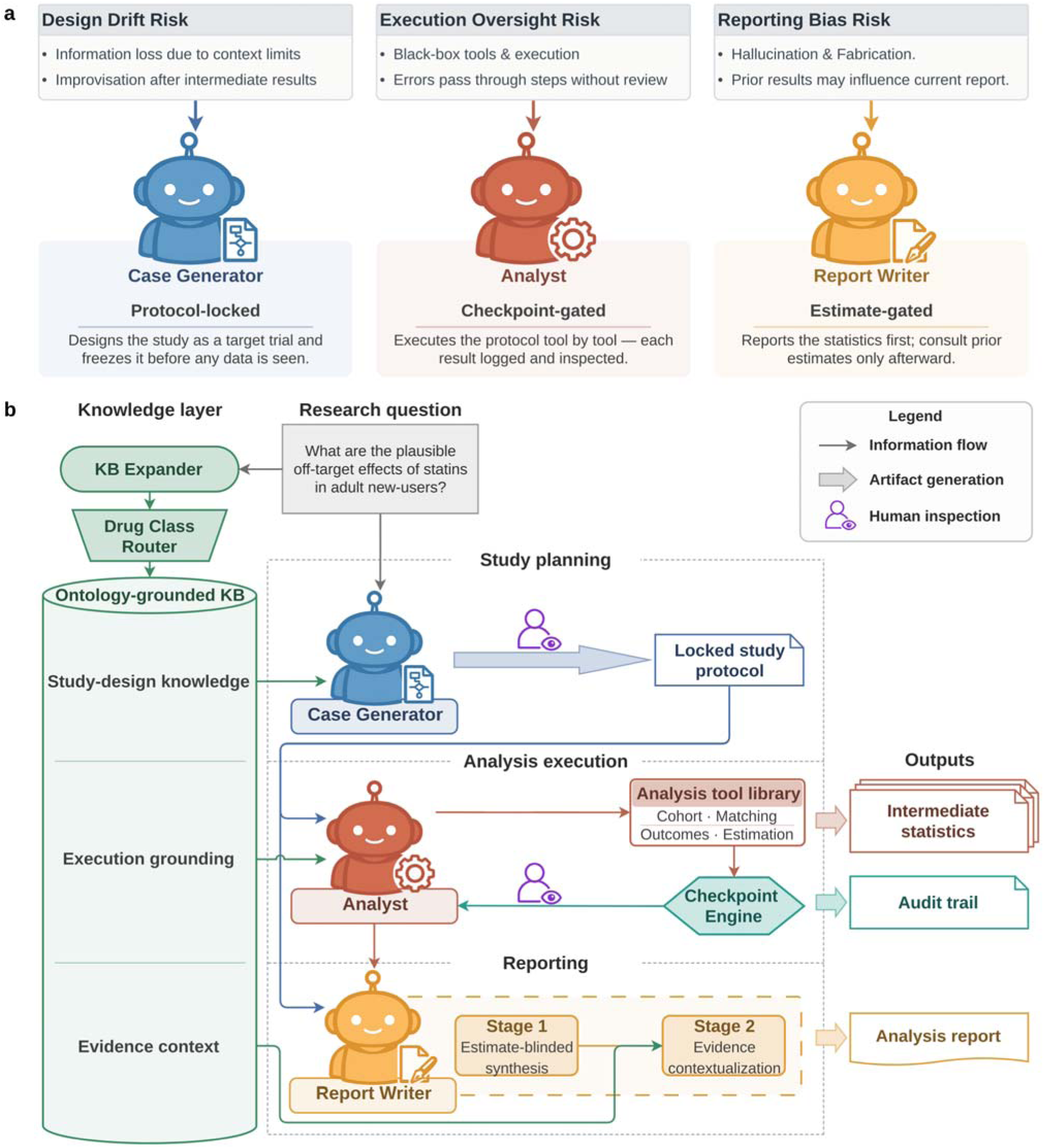
Agent roles and workflow in MedCheckpoint. a, The Case Generator translates natural language questions into structured study protocols. The Analyst coordinates analysis using human-verified deterministic tools that execute predefined statistical procedures. The Report Writer is instructed to present analytical findings before comparing them with prior evidence. b, Core study-design parameters are locked against modification by the Analyst. The knowledge base (KB) supplies source-traceable medical evidence for study design, execution, and interpretation. Checkpoints record execution status and govern whether analyses proceed. In human-in-the-loop mode, researchers can review decisions and intervene at checkpoints. Outputs include intermediate statistics, audit trails documenting analytical steps and decisions, and analysis reports.

MedCheckpoint uses a curated medical knowledge base to support this step. The knowledge base provides standardized drug and outcome definitions, as well as source-traceable medical evidence. These resources help the system identify both core safety outcomes, which are directly relevant to the question, and exploratory safety outcomes, which could reveal additional potential safety signals (Fig. 2).

**Fig. 2.**
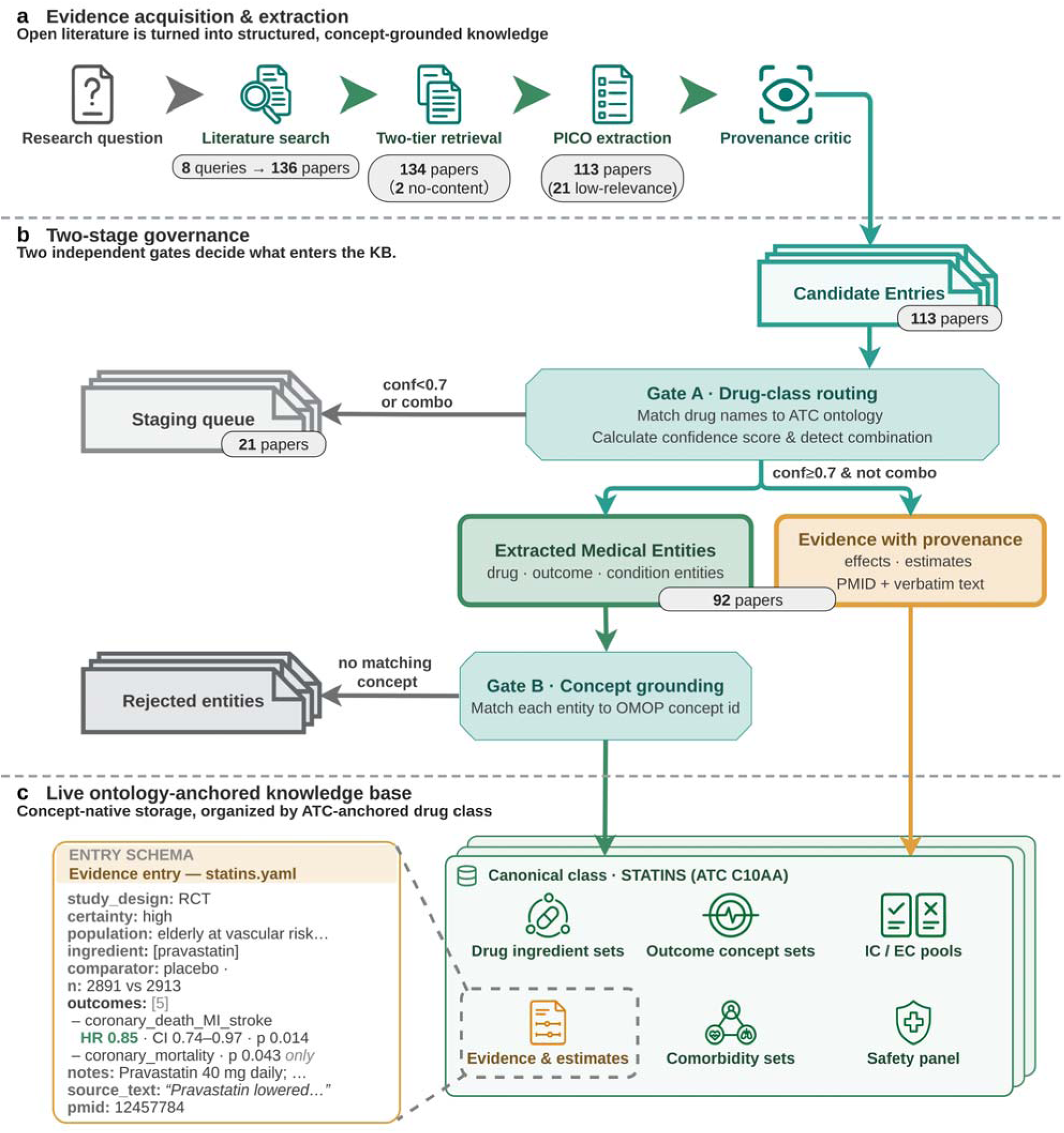
Medical knowledge base construction and organization. a, Literature retrieval prioritizes full text and uses abstracts when full text is unavailable. Structured extraction records study characteristics and findings. A separate source check compares the extracted information with the retrieved text. b, Drug-class routing assigns entries to standardized drug classes and holds uncertain or multi-class assignments for review. Concept grounding maps extracted medical entities to standard Observational Medical Outcomes Partnership (OMOP) concept identifiers. c, The knowledge base combines source-linked evidence, concept-based definitions, and shared comorbidity and safety-outcome resources. Paper counts represent one statin-related expansion run. The example entry summarizes the PROSPER trial^27^. KB, knowledge base; PICO, population, intervention, comparator, outcome; ATC, Anatomical Therapeutic Chemical; IC/EC, inclusion/exclusion criteria; conf, drug-class routing confidence; combo, labels spanning multiple drug classes; RCT, randomized controlled trial; HR, hazard ratio; CI, confidence interval (95% shown); MI, myocardial infarction; PMID, PubMed identifier.

After the protocol is generated, MedCheckpoint applies a protocol lock (Fig. 1b). This lock prevents the system from changing key study design elements during analysis. These locked elements include the exposure definition, outcome definitions, cohort criteria, and main statistical plan. The purpose of the lock is to reduce post hoc changes that could bias the analysis.

During the analysis stage, MedCheckpoint does not write new statistical code from scratch. Instead, it coordinates a set of human-verified deterministic tools. These tools perform cohort construction, confounding control, multi-outcome analysis, and statistical testing. This design makes the analytical workflow more reproducible and easier to audit.

MedCheckpoint also uses checkpoints during execution. These checkpoints summarize whether the planned analysis is feasible and whether each analytical step has been completed successfully. For example, a checkpoint can report whether the exposed cohort size has enough power, whether outcomes are observed frequently enough, or whether statistical models have converged. In fully automated mode, predefined rules stop analyses that fail critical feasibility checks. In human-in-the-loop mode, researchers can review checkpoint diagnostics, decide whether the analysis should continue, and provide feedback if needed.

In the reporting stage, MedCheckpoint first summarizes the results from the EHR analysis. It then compares these findings with evidence from medical literature. The final outputs include the study protocol, analysis results, checkpoint records, and the rationale for key decisions. These outputs are designed to support researcher review and auditability.

### Benchmark construction

We evaluated MedCheckpoint using a purpose-built pharmacovigilance benchmark of 54 natural language drug safety questions. The complete set of benchmark questions is provided in Supplementary Table 1. The benchmark was designed to test two capabilities. First, we assessed whether each system could produce appropriate epidemiological analyses. Second, we assessed whether each system avoided unsupported conclusions when the available data were insufficient.

The benchmark used a simulation-based longitudinal EHR dataset generated by Synthea, a patient-record simulator^12^. The dataset contains drug exposure and health outcome records for 2,709,803 individuals. Associations in these records can reflect assumptions of the data-generating model rather than true clinical drug effects.

We deliberately used simulated longitudinal EHRs to evaluate analytical rigor without treating reproduction of established drug effects as the criterion for success. This setting allowed us to examine whether agents remained faithful to the analyzed data when findings diverged from clinical expectations. This design was also intended to test whether agents could recognize potentially misleading signals and avoid treating apparent associations as established drug effects. Both capabilities are essential for drawing defensible conclusions in pharmacovigilance.

The 54 questions covered three types of pharmacovigilance tasks: 21 questions examined potential adverse drug reactions or off-target effects; 14 questions compared safety outcomes between drugs; and 19 questions evaluated dose-specific or patient subgroup-specific safety patterns.

### Baseline systems

We compared MedCheckpoint with two baseline systems. Baseline A was a general-purpose autonomous coding agent. It could write and execute code to answer the safety questions, but it did not have access to MedCheckpoint’s knowledge base, protocol lock, checkpoint rules, or specialized analysis tools.

Baseline B used the same execution framework as Baseline A, but it received additional methodological guidance derived from MedCheckpoint. This guidance was provided through prompts. The full study-specific prompts for both baselines are provided in Supplementary Note 1. Baseline B therefore tested whether prompting alone could reproduce the benefits of the full MedCheckpoint system.

All three systems used the same underlying LLM and the same EHR dataset. Each system was run three times for each question. The comparison with Baseline B was intended to isolate the added value of MedCheckpoint’s system design, including its curated knowledge base, deterministic tools, protocol lock, and checkpoint enforcement mechanisms. A qualitative comparison of system capabilities is provided in Supplementary Fig. 1.

### Overall quality and consistency of pharmacovigilance reports

The overall quality of analytical outputs and the methodological rigor of the analytical process are central to the value of autonomous EHR-based pharmacovigilance. Broader exploration of drugs and outcomes is useful only if the findings provide a sound basis for assessing potential safety signals and planning further investigation.

We therefore evaluated analytical quality using the completed reports and structured results from all three systems. The evaluation covered five dimensions of study design, analysis, and interpretation. Kimi-K3 (Kimi), DeepSeek-V4-Flash (DeepSeek), and MiniMax-M3 (MiniMax) served as LLM judges. Each scored every report on a 1–5 scale for each dimension. The scoring rubric described poor, intermediate, and strong performance in each dimension (Supplementary Note 2).

**Design Alignment** assessed whether the exposure definition, patient population, comparison group, and planned analysis addressed the research question. It also assessed whether reports justified these choices and the feasibility of the analysis.

**Cohort Validity** assessed whether reports accurately described the study groups and design, including how well matching made the groups comparable. It also assessed whether reports clarified the population to which findings applied and addressed residual confounding—bias from factors not fully controlled in the analysis.

**Statistical Rigor** assessed whether methods were appropriate and special cases, such as analyses with few outcome events, were handled correctly. It also assessed whether conclusions were supported by formal tests, estimates of uncertainty, and correction for multiple comparisons to limit false-positive findings.

**Outcome Validity** assessed whether outcome identification and interpretation were supported by the available data. This included handling outcomes that could not be measured reliably, assessing the validity of proxy measures used as indirect indicators, and comparing findings with prior evidence.

**Interpretive Integrity** assesses whether reports distinguish association from causation. It also assesses whether key claims are traceable to supporting evidence and whether residual confounding and uncertainty are explained for specific findings.

MedCheckpoint received the highest mean scores across all five dimensions from each of the three judges (Fig. 3a; Supplementary Table 2). The quality evaluation included 130 completed reports from MedCheckpoint, 158 from Baseline B, and 154 from Baseline A. Although MedCheckpoint completed fewer analyses, stopping before completion is warranted when the available data or study design cannot support a valid analysis. The following sections examine when analyses should proceed or stop and whether greater completion is accompanied by higher report quality.

**Fig. 3.**
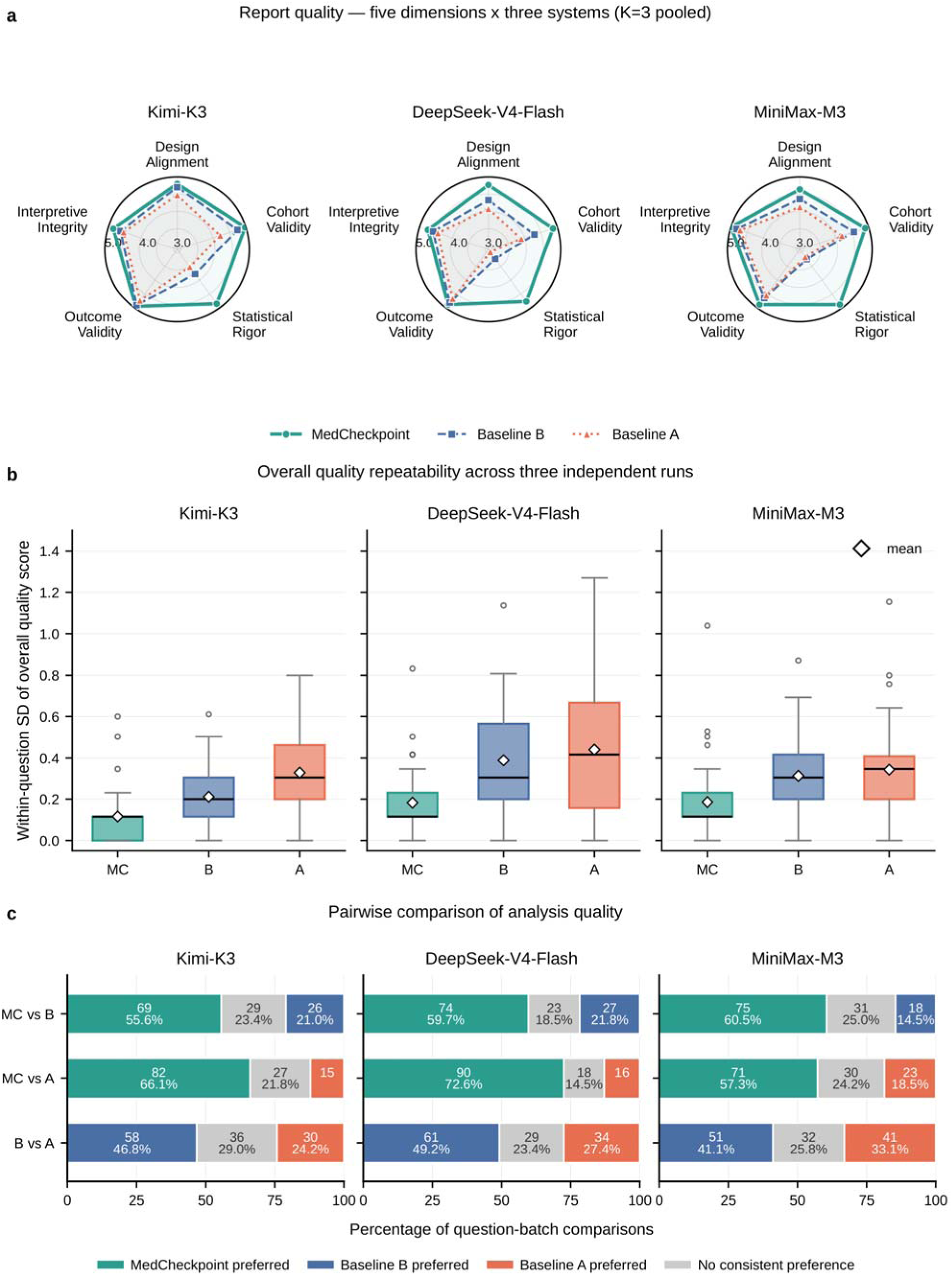
Report quality, performance consistency, and pairwise comparison. a, Mean scores for five quality dimensions, rated from 1 to 5; higher scores indicate better quality. Scores were pooled across three run batches (K=3). The sample comprised 130 MedCheckpoint (MC), 158 Baseline B (B), and 154 Baseline A (A) completed reports. Radar axes span 2.9–5. b, Within-question variation in report quality across three runs. Each run’s overall score is the mean of its five dimension scores. The sample standard deviation (SD) across runs measures variation; lower values indicate greater consistency. Only questions with three scored runs were included: 40 for MC, 50 for B, and 47 for A. Boxes span the 25th–75th percentiles, defining the interquartile range (IQR). Lines mark medians, diamonds means, and circles outliers. Whiskers extend to the most extreme observations within 1.5 IQRs of the box edges. c, Pairwise comparisons of 124 question–batch combinations with evaluable reports from all three systems. Each pair was judged in both presentation orders. Colored segments indicate preference for the same system in both orders. Gray denotes No consistent preference. It combines ties in both orders, a preference in one order with a tie in the other, and Winner flipped by order. The last category means the two judgments favored different systems.

In the order of the judges Kimi, DeepSeek, and MiniMax, MedCheckpoint’s overall mean scores across the five dimensions were 4.88, 4.81, and 4.86. The corresponding scores were 4.55, 4.25, and 4.32 for Baseline B and 4.29, 4.02, and 4.15 for Baseline A. With MiniMax-M3 held constant as the backbone LLM across all three systems, Baseline B scored above Baseline A but below MedCheckpoint. This pattern supports the added value of integrating knowledge support, analysis tools, and process constraints beyond methodological guidance alone. During development, MedCheckpoint generated reports of broadly similar quality when using either DeepSeek or MiniMax as its backbone LLM. This observation suggested that the framework’s methodological safeguards were not specific to a single backbone LLM.

**Design Alignment.** In the same judge order, MedCheckpoint’s mean scores were 4.80, 4.77, and 4.64. These scores are consistent with MedCheckpoint’s use of a study protocol to guide autonomous analysis. The protocol links the research question to key constraints on exposure, population, and comparison.

**Cohort Validity.** MedCheckpoint’s mean scores were 4.98, 4.87, and 4.90, all higher than those of both baselines. For example, in a report comparing clopidogrel with warfarin, MedCheckpoint specified that its estimates applied to treated patients within the region of propensity score overlap. The report also discussed channeling bias, or confounding when clinical characteristics influence treatment selection, and unmeasured clinical differences. In contrast, a separate Baseline A report on calcium channel blockers labeled a comparison with never-users as an active-comparator design. Its comparison group had no recorded use of calcium channel blockers, rather than being defined by use of an alternative treatment.

**Statistical Rigor.** This dimension showed the largest gap between MedCheckpoint and both baselines. MedCheckpoint’s mean scores were 4.85, 4.77, and 4.88. The corresponding scores were 3.78, 3.23, and 3.25 for Baseline B and 3.53, 2.99, and 3.17 for Baseline A.

MedCheckpoint’s human-verified deterministic tools correct for multiple comparisons and perform formal interaction tests, which assess whether associations differ across patient subgroups. The system requires the corresponding outputs before report generation, reducing reliance on agents to implement these steps during execution.

For example, MedCheckpoint’s exploratory statin report identified an association with proteinuria, or protein in the urine, that was statistically significant before correction. The report noted that the association did not remain significant after false discovery rate correction. This procedure limits the expected proportion of false-positive findings among results declared significant. By contrast, a separate Baseline A report on a stratified statin= analysis drew conclusions about differences in associations between subgroups from subgroup estimates alone. It did not report a formal interaction test.

**Outcome Validity.** MedCheckpoint’s mean scores were 4.95, 4.88, and 4.88. These higher scores reflect the reports’ distinction between associations that could be estimated and outcome claims unsupported by the available data. This distinction helps identify unanswered drug safety questions. It also helps avoid interpreting missing records or an inability to estimate an association as evidence that a risk is absent.

**Interpretive Integrity.** MedCheckpoint’s mean scores were 4.85, 4.75, and 4.99. Its reports first present findings from the analysis outputs, then separately compare them with evidence from the literature. This separation is consistent with the higher scores and helps researchers trace claims to their supporting evidence. It also helps distinguish findings in the study data from medical interpretations and judgments that require further validation.

**Performance consistency.** We next examined how much report quality scores varied when each system answered the same question three times. We measured this variation using the sample standard deviation (SD), with a lower SD indicating more consistent scores.

For each judge and system, we first averaged the five dimension scores within each run. We then calculated the SD of the three overall scores for each question. These question-level SDs were averaged across questions. We also calculated within-question SDs for each individual dimension and averaged them across questions. Only questions with valid scores in all three runs were included: 40 for MedCheckpoint, 50 for Baseline B, and 47 for Baseline A.

MedCheckpoint had the lowest mean within-question SD in overall quality for all three judges (Fig. 3b; Supplementary Table 3). In the order Kimi, DeepSeek, and MiniMax, its values were 0.116, 0.183, and 0.186. The corresponding values were 0.211, 0.389, and 0.313 for Baseline B and 0.329, 0.440, and 0.343 for Baseline A.

This pattern held across all 15 combinations of three judges and five quality dimensions. For Cohort Validity, MedCheckpoint’s mean within-question SDs were 0.029, 0.159, and 0.130. The corresponding values were 0.384, 0.657, and 0.470 for Baseline B and 0.551, 0.815, and 0.565 for Baseline A (Supplementary Fig. 2; Supplementary Table 4).

Because the eligible question sets differed across systems, we repeated the comparison using a common subset. This comprised 35 questions for which all three systems completed three runs and received valid scores. MedCheckpoint again had the lowest mean within-question SD for all three judges (Supplementary Table 3).

**Overall report comparison.** We also examined whether MedCheckpoint’s advantage in dimension-level scoring was evident when reports were compared as a whole. The analysis included 124 sets of completed reports, each addressing the same question in the same run batch. Each set contained one evaluable report from each system. Kimi, DeepSeek, and MiniMax compared report pairs presented under anonymous system labels. The dedicated prompt retained the five quality aspects but did not specify standards for assigning numerical scores. Judges were asked to select the better report or declare a tie, without assigning dimension scores. Each decision required supporting evidence from both reports. To assess sensitivity to presentation order, each pair was judged twice, with each report presented first once. In the same judge order, MedCheckpoint was preferred in both orders for 55.6%, 59.7%, and 60.5% of pairs against Baseline B. Against Baseline A, the corresponding proportions were 66.1%, 72.6%, and 57.3%. The preferred system changed between orders in 12.9%–22.6% of pairs in comparisons involving MedCheckpoint, across the three judges and two baselines (Fig. 3c; Supplementary Table 5). Nevertheless, each judge preferred MedCheckpoint over each baseline in both orders for more than half of the pairs.

**Inter-judge agreement.** Our automated, LLM-based evaluation was designed to scale with MedCheckpoint’s capacity to conduct analyses and generate reports on large-scale datasets. To assess the reliability of the automated ratings and their sensitivity to judge choice, we quantified agreement among three LLM judges using a common rubric. This experiment provided context for interpreting report quality and its consistency across repeated runs. In the pointwise evaluation of 442 completed reports, overall quality scores showed moderate agreement. The absolute-agreement intraclass correlation coefficient, ICC(A,1), was 0.625. The consistency coefficient, ICC(C,1), which allows for systematic differences in judge severity, was 0.657 (Supplementary Table 6).

As the systems’ backbone LLM, MiniMax supported analytical execution and report generation. As a judge, MiniMax used a dedicated rubric-based prompt requiring structured scores and justifications grounded in the submitted materials. Its ICC(A,1) values with Kimi and DeepSeek were 0.625 and 0.613, respectively, compared with 0.636 between Kimi and DeepSeek (Supplementary Table 7). For overall report quality, MiniMax’s agreement with the other judges was similar in magnitude to their agreement with each other.

Agreement varied across quality dimensions. Krippendorff’s ordinal α, which summarizes agreement among all three judges, was highest for Statistical Rigor, at 0.726. This dimension also had the highest linearly weighted Cohen’s κ for every judge pair, with values ranging from 0.535 to 0.611 (Supplementary Tables 8, 9). It was the same dimension in which MedCheckpoint showed the largest quality advantage over both baselines.

Across the five dimensions, all three judges’ scores differed by no more than one point for 87.8%–97.5% of reports. Scores in some dimensions were concentrated near the upper end of the scale. Both α and κ adjust for agreement expected by chance, which depends on the score distribution. We also report Gwet’s AC2, a prespecified complementary measure that uses a different chance-agreement model. Three-judge AC2 values ranged from 0.742 to 0.904 (Supplementary Tables 8, 9).

### Report safety under data limitations

In addition to quality and precision, safety and reliability are important aspects of pharmacovigilance reports. When data are limited, reports should state the limitations and explain how they affect the conclusions. They should not present overly confident findings.

We used a targeted challenge set of seven prespecified trap questions to assess whether systems could recognize and appropriately handle known data limitations (Fig. 4; Supplementary Note 3). Three questions involved use of both study drugs, which prevented the intended comparison between groups receiving one drug but not the other. Four involved key outcomes that could not be reliably identified from the available records. Each system was run three times per question, yielding 21 runs per system.

**Fig. 4.**
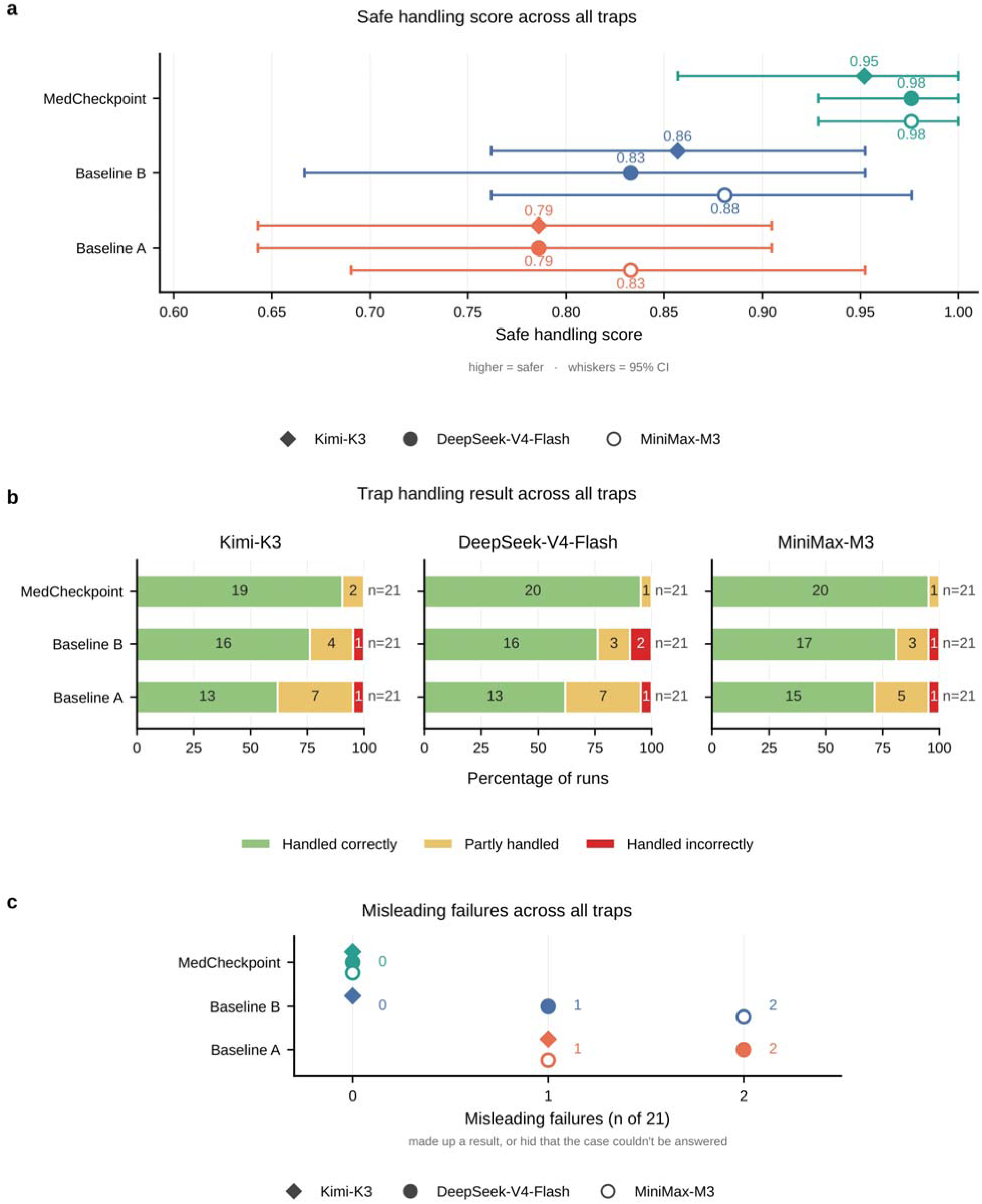
Report safety under data limitations. Seven questions tested data limitations. Three involved comparisons for which the intended patient groups could not be formed. Four involved key outcomes that could not be reliably identified from the available records. Each system was run three times per question, yielding 21 runs. a, Mean safe handling scores on a 0–1 scale. Handled correctly, Partly handled, and Handled incorrectly received scores of 1, 0.5, and 0, respectively. Scores were averaged across runs; higher values indicate better handling of data limitations. Whiskers show 95% confidence intervals from a question-level cluster bootstrap. This repeatedly resamples questions while keeping all runs of each sampled question together. b, Distribution of handling categories. Bar segments show percentages of runs, and labels give run counts. c, Counts of misleading failures, including unsupported findings or failure to disclose critical data limitations.

Kimi, DeepSeek, and MiniMax evaluated the resulting reports or abort records using a dedicated prompt and reference information on the known data limitations provided only to the judges. Using a prespecified rubric, they assigned one of three handling categories based on evidence in the system outputs.

**Handled correctly** required identifying the specific limitation and responding appropriately. For infeasible comparisons, this meant recognizing that the intended groups could not be formed. Systems then had to stop for that reason or explicitly state that the comparison was infeasible. For unmeasurable outcomes, systems had to identify the affected outcomes based on data coverage, avoid unsupported effect claims, and analyze measurable components.

**Partly handled** included noting confounding or group imbalance without recognizing that the comparison was infeasible. It also included recognizing that some outcomes were unmeasurable but incorrectly extending this limitation to measurable outcomes or downplaying its importance. Interpreting zero recorded events as evidence of no clinical effect also qualified. Vague or unrelated reasons for stopping also precluded a rating of Handled correctly.

**Handled incorrectly** included presenting infeasible comparisons as valid, reporting unsupported findings for unmeasurable outcomes, or omitting key limitations. It also included using clinically inappropriate proxies, meaning recorded outcomes that did not validly represent the outcomes of interest.

We separately counted misleading failures using five predefined error types. These included unsupported findings or effect claims for unmeasurable outcomes and clinically inappropriate proxies. They also included presenting infeasible comparisons as valid and omitting key unmeasurable outcomes or their measurement limitations. The fifth type was interpreting inadequate data coverage as evidence of no clinical association or risk. For each judge, runs were counted by their primary error type, separately from the handling categories.

None of the three judges identified misleading failures in MedCheckpoint’s 21 evaluated runs. In the order Kimi, DeepSeek, and MiniMax, Baseline B had 0, 1, and 2 such runs. Baseline A had 1, 2, and 1, respectively (Fig. 4c).

Two examples illustrate the distinction between answering from clinical knowledge and drawing research conclusions from the specified data. Baseline A answered a question about sex differences in amlodipine-associated peripheral edema, or swelling caused by fluid accumulation in the limbs. It reported incidences of 5.6% in men and 14.6% in women, describing these as approximate figures from prescribing information. It did not inspect the data or disclose that peripheral edema was not reliably measurable in the dataset. All three judges counted this response as a misleading failure.

Baseline B answered a metformin question involving patients with chronic kidney disease without inspecting the data. It discussed lactic acidosis, in which lactate accumulation is accompanied by increased blood acidity. It also described kidney function thresholds for metformin use and vitamin B12 deficiency. The response did not disclose that lactic acidosis and B12 deficiency were not reliably measurable in the dataset. All three judges classified this run as Handled incorrectly, but only MiniMax included it in Misleading failures. The original execution records likewise showed that both responses omitted these data-coverage limitations.

MedCheckpoint also had the most runs classified as Handled correctly. In the order Kimi, DeepSeek, and MiniMax, the counts were 19, 20, and 20 of 21 runs. The corresponding counts were 16, 16, and 17 for Baseline B and 13, 13, and 15 for Baseline A. All remaining MedCheckpoint runs were Partly handled; none were Handled incorrectly (Fig. 4b).

To summarize handling performance, we assigned scores of 1 to Handled correctly, 0.5 to Partly handled, and 0 to Handled incorrectly. The Safe handling score was the mean across runs, with higher values indicating better handling of data limitations. MedCheckpoint had the highest scores: 0.952, 0.976, and 0.976. The corresponding scores were 0.857, 0.833, and 0.881 for Baseline B and 0.786, 0.786, and 0.833 for Baseline A (Fig. 4a; Supplementary Table 10).

The largest differences in correct handling occurred when the intended comparison was infeasible. In all 9 runs of this type, MedCheckpoint identified the limitation and terminated the analysis, and all three judges rated them as Handled correctly. The corresponding counts were 6, 5, and 6 for Baseline B and 2, 3, and 4 for Baseline A. In runs not rated Handled correctly, both baselines often noted confounding or group imbalance without recognizing that the intended comparison was infeasible.

For the 12 runs involving key unmeasurable outcomes, MedCheckpoint instead generated reports in every run, with 10, 11, and 11 classified as Handled correctly. The remaining runs were Partly handled because judges considered some outcomes labeled unmeasurable to be measurable in these data. MedCheckpoint thus aborted infeasible comparisons while continuing analysis and reporting when measurable components remained. Safe handling scores by limitation type are shown in Supplementary Fig. 3.

### Analysis completion and termination across systems

Analysis completion varied across systems and repeated runs of the same question. We examined these differences and whether completing more analyses was accompanied by higher report quality.

We selected questions retrospectively, using the existing execution records and initial classifications of benchmark outputs. Questions from the preceding data-limitation tests were excluded. A question was included if at least one system had a run initially classified as not completed. We then included all three runs from each system for every selected question.

This yielded 14 questions and 42 runs per system (Supplementary Table 11). The resulting proportions describe system behavior within this selected sample, rather than completion rates across the full benchmark.

We used LLM assistance to organize the original reports, analysis outputs, and execution logs. Final judgments were made through human review. We classified execution outcomes into three categories, separately from report quality. Completed with available data denotes conducting a data analysis and reporting findings and limitations based on its outputs. Properly stopped (stopped at cohort or matching checks) denotes stopping when cohort or matching checks indicate that analysis cannot proceed under the current protocol. These runs stop before subsequent analysis and reporting. Fabricated answer without analyzing denotes answering the research question without analyzing the specified data, including substituting general medical knowledge for data analysis.

Of Baseline B’s 42 runs, 40 were classified as Completed with available data and 2 as Fabricated answer without analyzing. Baseline A had 36 and 6 runs in these categories, respectively. All 8 baseline runs without completed data analyses substituted medical knowledge or literature-based answers for analysis of the specified data.

Of MedCheckpoint’s 42 runs, 20 were classified as Completed with available data and 22 as Properly stopped (stopped at cohort or matching checks). These stopped runs explicitly reported their status. No MedCheckpoint runs were classified as Fabricated answer without analyzing (Fig. 5; Supplementary Table 12).

**Fig. 5.**
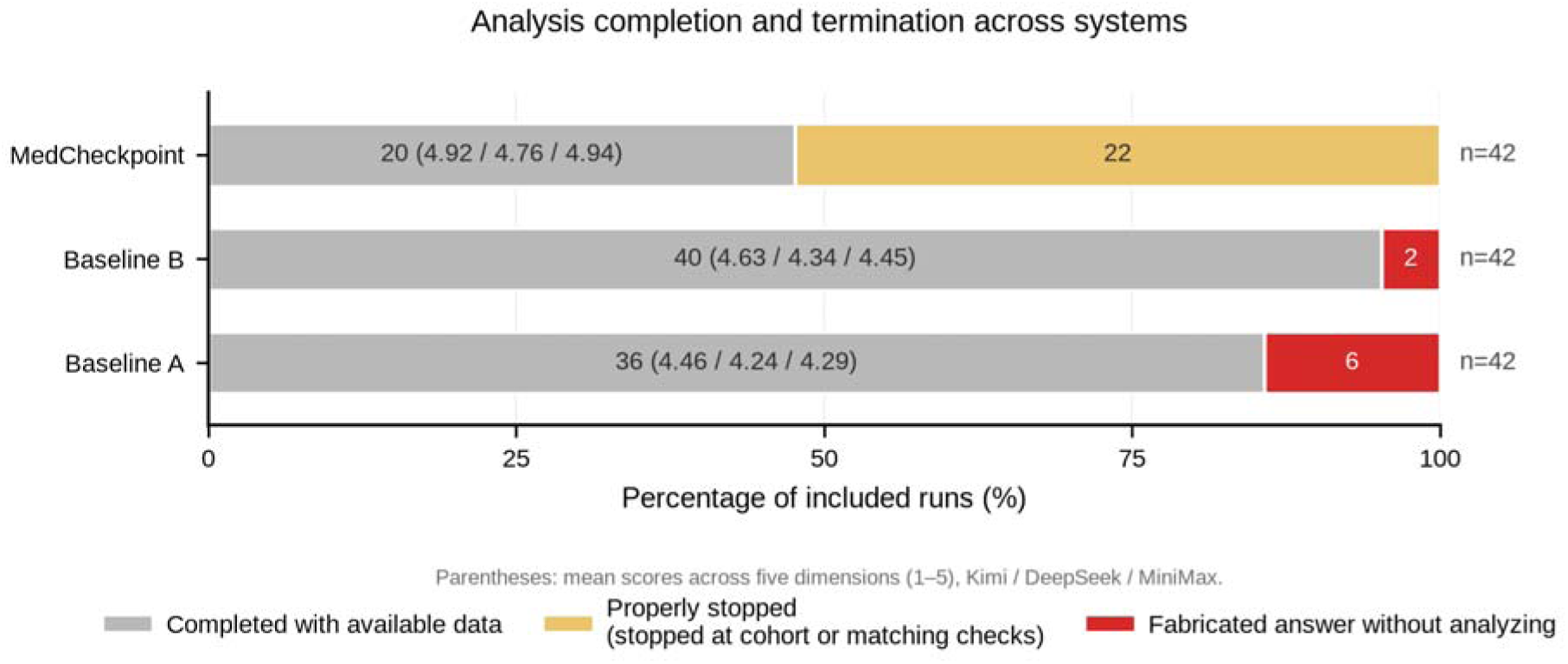
Analysis completion and termination across systems. Bars summarize a retrospective audit of 14 questions. Questions from the data-limitation tests were excluded. A question was included if at least one system had a run initially classified as not completed. All three runs from each system were included for every selected question, yielding 42 runs per system. Segment widths show percentages within this selected audit sample. Labels give run counts. Parentheses in the gray segments give mean quality scores for completed reports on a 1–5 scale. The five quality dimensions are weighted equally. Scores are listed in the order Kimi, DeepSeek, and MiniMax.

Both baselines were instructed to base answers on the data and state analytical limitations, but lacked execution gates enforcing these requirements. MedCheckpoint requires the necessary analysis outputs before a formal report can be generated. If cohort or matching checks trigger an abort, its checkpoint engine blocks further analysis and reporting tool calls.

In MedCheckpoint, completion varied across runs of the same question. This variation arose partly from additional criteria for including or excluding patients in independently generated protocols. Some criteria prevented a study group from being formed. Runs without those criteria could still complete exploratory analyses. Their findings nevertheless required caution because of limitations in matching quality and residual confounding.

Some aborts could therefore represent missed opportunities for feasible analysis. Saved protocols and records explaining why analyses stopped help researchers identify design criteria that could be adjusted. Researchers can then provide targeted methodological guidance to support feasible exploratory analyses in subsequent runs.

We next compared the quality of completed reports in this sample, reusing their earlier scores on the five-dimension rubric and 1–5 scale. Overall quality was calculated by averaging the five dimension scores, giving each dimension equal weight. In the order Kimi, DeepSeek, and MiniMax, MedCheckpoint’s overall mean scores were 4.92, 4.76, and 4.94. All were higher than those of both baselines (Fig. 5; Supplementary Table 13).

To assess whether differences in completed tasks influenced this comparison, we also examined analyses completed by all three systems. We grouped their reports by question and run batch, with one report from each system per group. This yielded 15 groups covering 10 questions, with 15 reports per system. MedCheckpoint again had the highest overall mean scores for all three judges. Statistical Rigor showed the largest gap in all six comparisons across the three judges and two baselines. MedCheckpoint scored 0.93–1.53 points higher in this dimension (Supplementary Table 14).

The higher completion counts of the two baselines did not translate into higher report quality. This pattern is consistent with MedCheckpoint’s use of human-verified deterministic statistical tools. These tools estimate associations, perform formal tests, and correct for multiple comparisons.

### A case study of researcher-guided protocol refinement

Human expertise remains a cornerstone of clinical research. MedCheckpoint enables researchers to guide study design and intervene at analytical checkpoints. These review points allow timely input of domain knowledge to guide the analysis and correct its course when needed.

We present a demonstration of researcher-guided protocol refinement after automated runs encountered design-related barriers. The selected question compared amlodipine and metoprolol in adults starting treatment for hypertension, or high blood pressure. It covered cardiovascular and adverse outcomes. We compared two conditions, without and with guidance. The unguided condition used only the original research question. In the guided condition, a researcher note informed by cohort construction and abort records was added to the question before a new protocol was generated.

Both conditions used the same system version, underlying LLM, and study dataset. We conducted five independent runs per condition. Each run generated a new protocol and then proceeded in fully automated mode. Completion required both an analysis report and a final results table. We reviewed locked protocols, cohort and matching outputs, and abort records to assess how guidance informed study design and execution.

In all five unguided runs, the Case Generator required hypertension for inclusion. Four protocols added exclusions for varying combinations of other conditions. These included heart failure, in which the heart cannot fill or pump blood adequately, and prior myocardial infarction (heart attack). Two runs stopped because the metoprolol comparison group was empty. Two others stopped because the amlodipine group was empty. None of these four runs reached statistical analysis.

The only completed run omitted these additional exclusions. It included 48,025 amlodipine users and 35,118 metoprolol users. Matching yielded 3,454 pairs, each containing one patient from each drug group. This run showed that an exploratory comparison remained feasible when the population with hypertension was retained without these additional exclusions.

Based on the cohort construction and abort records, the researcher provided the following researcher note:

Keep the study focused on its intended population — adults with hypertension who are new users of amlodipine or metoprolol. Within that hypertensive population, do not narrow the cohort further by excluding patients with common cardiovascular comorbidities such as heart failure, prior myocardial infarction, atrial fibrillation, or stroke: because metoprolol is frequently prescribed to such patients, excluding these conditions removes most of the comparator arm and makes the comparison infeasible. Retain hypertension as the core eligibility requirement, and keep an analyzable population representative of real-world users of both drugs, unless a specific exclusion is essential to the research question or patient safety.

The note addressed which patients to include or exclude. It did not specify an expected effect direction or magnitude. The same note was used in all five guided runs.

With guidance, all five runs retained hypertension as an inclusion criterion and omitted the additional cardiovascular exclusions. Completion increased from 1/5 without guidance to 5/5 with guidance. Cohort sizes were identical to those of the completed unguided run. Matching yielded 3,448–3,454 pairs per run.

The reports nevertheless noted that retaining patients with coexisting cardiovascular conditions could leave residual confounding despite propensity score adjustment. Patients receiving the two drugs could still differ in their reasons for treatment and disease severity. The matched groups also differed in the year patients entered the study and the frequency of visits and tests. These differences could affect whether outcomes were recorded or detected. The reports therefore treated the findings as exploratory associations requiring further validation, not as a basis for clinical prescribing recommendations.

This demonstration illustrates how saved protocols and abort records can support researcher-guided protocol refinement. The researcher used these records to identify restrictive eligibility criteria and provide natural language guidance for subsequent protocols. MedCheckpoint then executed the new protocols, performed statistical analyses, and reported results, allowing exploratory analyses to proceed.

### A case study of stratified pharmacovigilance analysis

We present a case study of MedCheckpoint’s application to stratified pharmacovigilance analysis. The case addresses the question: “Is the hyperglycemic effect of systemic corticosteroids modified by pre-existing diabetes?” Systemic corticosteroids act throughout the body. Hyperglycemia means abnormally high blood glucose.

Answering this question requires retaining patients with and without diabetes before treatment. Stratified analyses estimate the drug–hyperglycemia association separately in these two groups. A formal interaction test assesses whether these associations differ. MedCheckpoint, Baseline A, and Baseline B attempted stratified analyses but differed in how they revised analyses, handled few outcome events, and interpreted evidence (Fig. 6). The simulation-based setting also allowed us to examine how systems responded when observed associations conflicted with established clinical expectations.

**Fig. 6.**
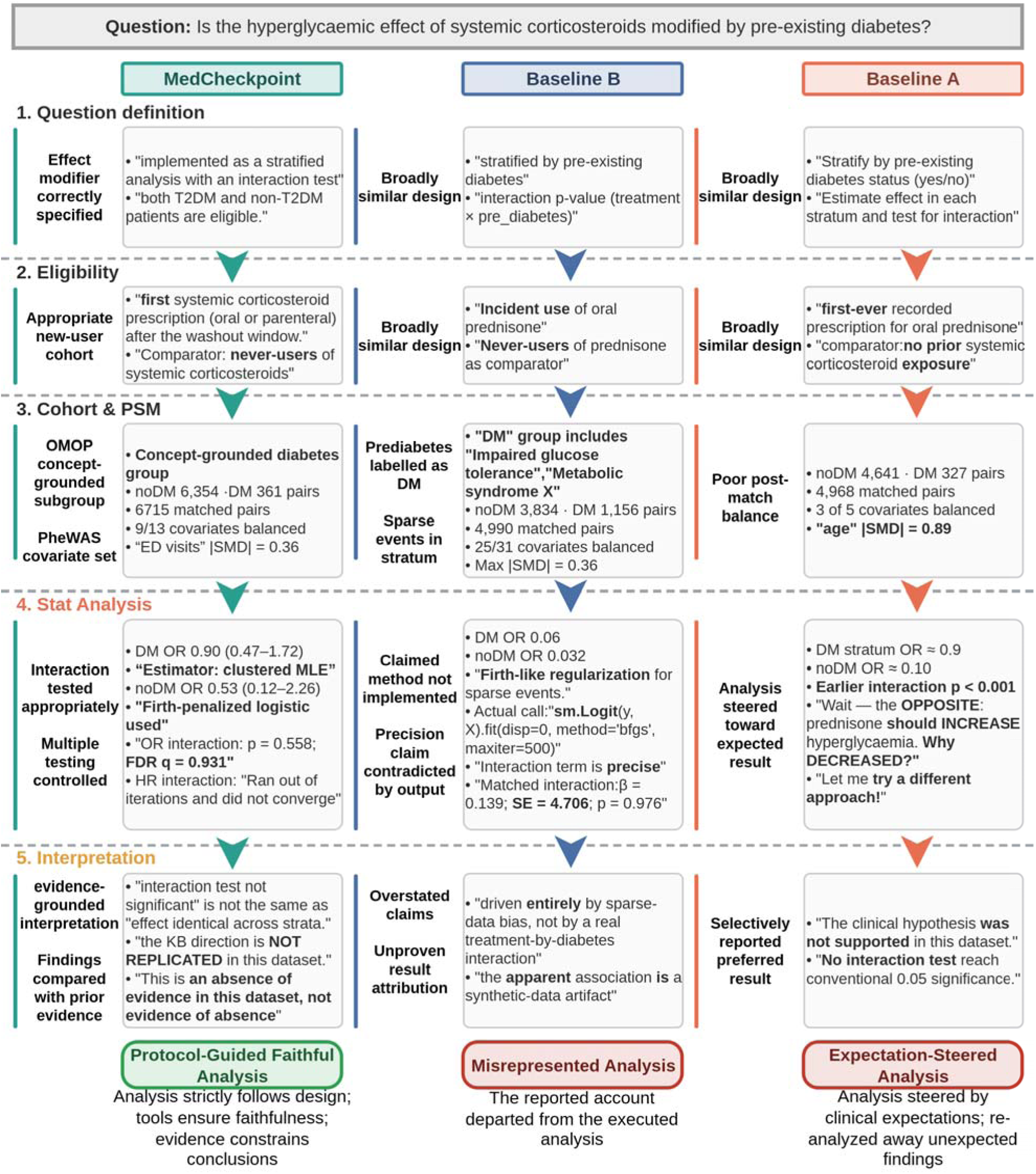
A case study of stratified pharmacovigilance analysis. Comparison of study design, execution, and interpretation for the corticosteroid–hyperglycemia analysis using simulated longitudinal electronic health record data. Analyses are performed separately in patients with and without pre-existing diabetes. Subgroup definitions and outcome windows differ across systems. Boxes contain excerpts and analytical summaries. Adjacent annotations are the authors’ assessments. DM and noDM denote each system’s groups with and without pre-existing diabetes, respectively. T2DM denotes type 2 diabetes mellitus. A covariate is counted as balanced when its absolute standardized mean difference (SMD) is below 0.1. OR ranges are 95% confidence intervals. β and SE denote the interaction coefficient and its standard error, respectively. Both are on the log-odds scale used in logistic regression. PSM, propensity score matching; OR, odds ratio; HR, hazard ratio; FDR, false discovery rate; MLE, maximum likelihood estimation; OMOP, Observational Medical Outcomes Partnership; PheWAS, phenome-wide association study; ED, emergency department; KB, knowledge base.

Baseline A compared new users of oral prednisone with never-users. Initial propensity score matching yielded 4,968 pairs, but absolute standardized mean differences (SMDs) for age were 0.764 without pre-existing diabetes and 0.894 with diabetes. This indicates substantial imbalance. Baseline A discarded these matches and analyzed the full cohort of 214,366 individuals. It used regression adjustment to account for measured patient characteristics in the statistical model.

Baseline A’s early subgroup model omitted the exposure variable identifying prednisone use. It misinterpreted the estimated age association as the treatment association, then corrected this error. After correction, the odds ratio (OR) for hyperglycemia in the subgroup without pre-existing diabetes was 0.112, with a 95% confidence interval (CI) of 0.073–0.172.

Pooled logistic models combined both diabetes subgroups to test interaction. For hyperglycemia diagnoses recorded at any time after cohort entry, the interaction p value was 4.02 × 10 ¹. The composite outcome counted either hyperglycemia or newly initiated diabetes medication within 365 days. Its interaction p value was 7.34 × 10.

Baseline A’s execution log then questioned the protective association: “Prednisone should INCREASE hyperglycemia. Why does it show DECREASED?” Baseline A cited selection bias and population differences as reasons for reanalysis. It argued that the groups differed in ways that could distort the comparison. It required at least one visit in the year before cohort entry in both groups and redesigned matching.

When individual matching proved slow, Baseline A switched to matching within groups defined by diabetes status, sex, and age. Each treated patient could have up to five controls. After refitting, interaction p values were 0.27 for hyperglycemia diagnoses and 0.10 for the 365-day composite outcome. A subsequent Cox model, which analyzes time to an event, yielded an interaction p value of 0.34. Two of the five measured patient characteristics (covariates) still had absolute SMDs above 0.1, with a maximum of 0.54.

The final report concluded that effect modification was unsupported. It did not present the corrected early results or explain the changed interaction evidence after revising the population and matching. Observed results and their conflict with clinical expectations thus influenced Baseline A’s subsequent analytical choices. However, the report did not adequately describe how these choices affected its conclusions.

Baseline B also analyzed oral prednisone, with hyperglycemia within 90 days as the primary outcome. However, its definition of pre-existing diabetes included impaired glucose tolerance and metabolic syndrome. Impaired glucose tolerance is an intermediate state of abnormal glucose regulation, not a diabetes diagnosis. Metabolic syndrome is a cluster of conditions associated with increased diabetes and cardiovascular risk. The matched cohorts of Baseline B and MedCheckpoint shared 4,687 treated individuals with identical treatment initiation dates. Among them, 881 received Baseline B’s diabetes label solely because of prior impaired glucose tolerance records. Its diabetes subgroup therefore extended beyond diagnosed diabetes.

Baseline B tried different propensity score models and matching calipers. Calipers limit the permitted difference between propensity scores for a match. Final matching produced 4,990 pair records from 4,990 treated individuals and 2,421 distinct controls, with controls reused. It reported both matched estimates and regression-adjusted estimates from the unmatched cohort. Six of 31 covariates remained imbalanced after matching, with absolute SMDs above 0.1 and a maximum of 0.36.

Under Baseline B’s subgroup definitions, the primary outcome occurred in 0 treated patients and 1 control without diabetes. The corresponding counts in the diabetes subgroup were 1 and 16. Baseline B considered Firth penalized likelihood, which reduces small-sample bias in logistic regression, or other approaches. However, the final script’s “Firth-like regularization” label did not describe its implementation. It used ordinary, unpenalized logistic regression with the Broyden–Fletcher–Goldfarb–Shanno optimizer. This algorithm fits the model but does not itself apply the Firth correction. Recognizing the problem and proposing an appropriate method did not ensure its implementation.

In Baseline B’s diabetes subgroup, the OR was 0.063 with a 95% CI of 0.008–0.474. Interaction p values were 0.91 in the unmatched adjusted analysis and 0.98 in the matched analysis. The report acknowledged that the subgroup without diabetes provided insufficient information for a reliable answer. Yet it attributed apparent subgroup differences entirely to sparse-data bias and described the interaction as “precisely estimated”. However, the matched interaction coefficient had a standard error of 4.71 on the logarithmic odds scale used by logistic regression. This large standard error indicated substantial uncertainty, contradicting the claim of precision.

The report also interpreted the protective association in the diabetes subgroup as a likely artifact of the data-generating process. However, failure to detect interaction with sparse data did not by itself establish that bias or artifacts caused the subgroup differences. Despite acknowledging limitations, Baseline B had not fully translated methodological guidance into reliable execution and interpretation.

MedCheckpoint followed the Case Generator’s protocol, with core study-design parameters fixed by protocol lock. The protocol specified pre-existing type 2 diabetes as the effect modifier and required retaining both subgroups. It compared adult new users of systemic corticosteroids with never-users over a two-year outcome window. Execution records confirmed that all patients in the exposed group started prednisone.

Matching was performed separately within the two diabetes subgroups. It yielded 6,715 one-to-one pairs without patient reuse: 6,354 without and 361 with pre-existing type 2 diabetes. After matching, 9 of 13 covariates had absolute SMDs below 0.1. Emergency care utilization remained imbalanced, with an absolute SMD of 0.357. After excluding pairs with unresolved diagnosis information, 6,674 pairs entered the hyperglycemia analysis. Matching diagnostics were retained for inspection.

MedCheckpoint’s Analyst uses human-verified deterministic statistical tools to handle sparse data, perform formal interaction tests, and correct for multiple comparisons. In patients without pre-existing type 2 diabetes, there were 2 hyperglycemia events in the exposed group and 4 in controls. The tool used Firth logistic regression, yielding an OR of 0.528 with a 95% CI of 0.123–2.260.

In the subgroup with pre-existing type 2 diabetes, it used ordinary logistic regression with standard errors clustered by matched pair. This accounts for dependence between patients in the same pair. The OR was 0.901 with a 95% CI of 0.471–1.724, and the overall OR was 0.725 with a 95% CI of 0.411–1.278.

The pooled logistic model tested the interaction between corticosteroid exposure and diabetes status. A likelihood-ratio test compared models with and without this interaction term, yielding p = 0.5575. Benjamini–Hochberg correction for multiple comparisons gave a q value of 0.9309.

The hazard ratio (HR) interaction model failed to converge to a stable numerical solution. The tool flagged this failure and returned no interaction p value. Structured outputs recorded the estimation methods, correction results, and failure reasons. These records allowed the actual statistical execution to be checked, rather than inferred from method names.

MedCheckpoint’s Report Writer stated that the OR test did not detect statistically significant effect modification. This did not establish equal associations across subgroups. The failed HR model provided no interaction test result, rather than evidence against effect modification.

The report also compared the findings with knowledge base evidence. The data did not reproduce the established increase in hyperglycemia risk with corticosteroids. The report described this as insufficient evidence in these data, not evidence of no association. Clinical expectations therefore informed interpretation without replacing the findings from the current analysis. This case illustrates how MedCheckpoint uses deterministic tools to execute statistical methods and make their implementation verifiable. Recorded estimates, corrections, and failures support interpretation consistent with the available evidence.

## Discussion

In this study, we present MedCheckpoint, a domain-grounded multi-agent framework for automated postmarketing pharmacovigilance. Our results show that LLM agents, when constrained by curated biomedical knowledge, prespecified epidemiologic rules, deterministic statistical workflows, and human-in-the-loop checkpoints, can generate more reliable and reproducible pharmacovigilance analyses than generalist coding-agent baselines. These findings suggest that LLM-based systems have the potential to help shift pharmacovigilance from a predominantly reactive, case-by-case paradigm toward a more systematic, scalable, and proactive model of postmarketing drug safety surveillance.

The potential public health value of such systems is particularly high for adverse effects that are delayed, uncommon, population-specific, or difficult to detect in preapproval trials. Randomized controlled trials are often underpowered to identify rare harms and can exclude older adults, patients with multimorbidity, pregnant individuals, or people receiving polypharmacy^1,13,14^. Large-scale electronic health record and claims data can complement trials by capturing medication use across broader and more clinically heterogeneous populations^15^. However, this opportunity has historically been constrained by the expertise and time required to translate each safety question into a valid observational study^16^. MedCheckpoint addresses this bottleneck by automating major components of the pharmacovigilance pipeline while preserving key safeguards required for epidemiologic validity. Although automated screening cannot by itself establish causality, it can prioritize signals for confirmatory evaluation and thereby reduce preventable harm from adverse drug reactions.

Our findings also highlight an important distinction between automation and rigor. General-purpose LLM agents are increasingly capable of writing code, querying databases, and producing analytical reports. However, in clinical epidemiology, plausible-looking outputs are insufficient. A system can generate syntactically correct code and fluent explanations while silently violating core design principles, such as incorrect cohort entry dates, immortal time bias, inappropriate comparator selection, outcome misclassification, omitted confounding control, or inconsistent reporting of statistical results. These failures are not merely technical errors; they can directly alter safety conclusions^17,18^. Therefore, in medical applications, the primary challenge is not only whether an LLM can automate a task, but whether it can do so under methodological constraints that are transparent, auditable, and reproducible.

MedCheckpoint was designed around this principle. Rather than relying solely on prompt instructions or unconstrained agent reasoning, the system grounds each analytic stage in a curated knowledge base and locks critical analytical decisions before execution. This design reflects the reality that different stages of pharmacovigilance require different forms of expertise. Query interpretation requires clinical and pharmacologic knowledge; cohort construction requires epidemiologic design rules; analysis execution requires deterministic and validated statistical procedures; and report generation requires conservative interpretation of uncertainty. By organizing the knowledge base around these stages, MedCheckpoint reduces the need for the LLM to improvise under uncertainty. The LLM is used primarily as an interface and coordinator, while high-risk analytical steps are governed by prespecified rules and human-verified computational tools.

This architecture provides a practical approach to making LLM agents both useful and conservative. In many biomedical settings, excessive model autonomy can be hazardous because the model can fill gaps with unsupported assumptions. Conversely, overly rigid systems can fail to generalize across diverse pharmacovigilance questions. MedCheckpoint attempts to balance these competing needs by allowing flexible natural-language interaction and task decomposition while constraining decisions that determine validity. The system can interpret a broad range of drug safety questions, but it must map them to standardized study designs, verified code modules, and predefined reporting structures. This separation between flexible reasoning and locked execution is critical for safety-critical artificial intelligence. It allows the system to benefit from LLM capabilities without allowing the model to freely invent methodology during analysis.

The quality and scope of the curated knowledge base are central to system performance. A knowledge base that is too narrow can fail to support diverse pharmacovigilance questions, whereas an overly broad or weakly curated knowledge base can introduce conflicting recommendations, irrelevant context, or outdated information. This raises an important design question for future LLM-based medical systems: whether knowledge should be curated prospectively, retrieved dynamically at query time, or combined through a hybrid approach. Prospective curation offers stability, auditability, and methodological consistency, which are particularly important for regulatory-grade evidence generation. Dynamic retrieval could improve coverage and currency, especially for newly approved drugs or emerging adverse event literature, but it also introduces risks of retrieving low-quality or inconsistent information.

Thus, this work still has some directions for future exploration. Future work should examine the optimal size, structure, updating frequency, and governance process for pharmacovigilance knowledge bases. A tiered model could be preferable, in which core epidemiologic principles and statistical workflows are fixed and version-controlled, while drug-specific clinical evidence is periodically updated through transparent review. Besides, with additional resources, independent assessments by pharmacovigilance experts could also be integrated into the evaluation framework. A key next step is to deploy MedCheckpoint in real-world research settings and evaluate its performance using routinely collected clinical EHRs.

## Methods

### Ethics statement

This study involved no human participants or protected health information. Review by an institutional review board was therefore not required.

### Data source and OMOP CDM preprocessing

We evaluated the systems using a simulation-based longitudinal EHR dataset generated by Synthea^12^. It contains records for 2,709,803 simulated individuals, describing their health and care over time. The data follow the Observational Medical Outcomes Partnership Common Data Model (OMOP CDM)^19^. This model organizes healthcare data into standardized tables for conditions, drug exposures, visits, measurements, and observation periods. Conditions are primarily coded using SNOMED CT, a standardized clinical terminology. Medications are primarily coded using RxNorm, a standardized drug terminology.

During preprocessing, we used the OMOP vocabulary to replace numeric concept identifiers with their corresponding names. Each table was stored in Parquet, a file format that organizes data by column. During analysis, MedCheckpoint loads data only when needed to reduce memory use.

We used these records to construct study cohorts, describe patients’ characteristics at study entry, and identify subsequent outcomes. The evaluation covered exploratory pharmacovigilance workflows from natural language questions to analyses of multiple outcomes. It assessed the quality of study design, analytical execution, and interpretation. We also examined whether systems recognized data limitations and avoided unsupported conclusions when comparisons or outcomes could not be evaluated.

The drugs and outcomes available for analysis depend on what Synthea simulates and which codes appear in the dataset. Some research questions therefore cannot be addressed. All individuals have an observation-period record specifying the time span covered by their data. Cohort construction checks whether this period meets study-specific duration requirements. Associations observed in the study data were not interpreted as evidence of drug safety in real-world populations.

### MedCheckpoint system overview

MedCheckpoint comprises three agents: the Case Generator, Analyst, and Report Writer (Fig. 1a). Together, they produce an observational analysis report from a natural language drug safety question.

The Case Generator translates the question into a structured protocol specifying the population, drug exposure, comparison group, and outcomes. It uses source-traceable medical evidence and standardized drug and outcome definitions from the knowledge base. Core outcomes include those targeted by the question, known drug effects, and predefined serious safety outcomes. Exploratory outcomes extend screening to other medically plausible outcomes.

The Analyst carries out the protocol by selecting and calling human-verified deterministic tools. These tools execute predefined calculations. A protocol lock restricts changes to core study-design parameters during execution. The checkpoint engine governs analytical execution and records key decisions. In human-in-the-loop mode, researchers can review intermediate results and intervene at these checkpoints.

The Report Writer prepares the report from the protocol and analysis outputs, including statistical estimates and diagnostic summaries. It is instructed to present the current analysis results before comparing them with evidence from the knowledge base. The report separates data-derived findings from literature-informed interpretation and describes uncertainty and study limitations.

Tool calling for all three agents is implemented with LangChain (version 1.3.9), and LLMs are accessed through a unified LiteLLM (version 1.89.0) interface. Backbone LLMs for knowledge-base construction and agent execution can be configured independently. A fixed random seed makes randomized steps in control sampling and matching repeatable for the same inputs. LLM decisions can still vary between runs. We therefore assess consistency in report quality scores through independent repeated runs.

### Knowledge base construction and concept grounding

MedCheckpoint organizes its medical knowledge base around a drug-class ontology (Fig. 2). This structured representation defines drug classes and their members using the Anatomical Therapeutic Chemical (ATC) classification. Each class is specified by a set of standard RxNorm concepts that identify its active ingredients.

For each drug class, evidence is organized using the population, intervention, comparator, and outcome (PICO) framework. This records who was studied, which treatments were compared, and which outcomes were assessed. The knowledge base also stores potential confounders, literature-derived patient inclusion and exclusion criteria, and source information. A shared library supplies outcome definitions for core and exploratory screening. A safety panel lists serious adverse events as core outcome candidates, regardless of drug class.

Knowledge expansion uses an LLM-based process separate from analysis execution. The system generates candidate drug classes and literature search queries from the research question. It searches PubMed and Semantic Scholar. It prioritizes full-text articles from PubMed Central and uses abstracts when full text is unavailable. An LLM extracts structured information from the available text. A separate LLM call then checks the extraction against the source text.

Candidate drug-class labels are matched to classes in the ontology. Entries meeting the acceptance criteria are added to the main knowledge base. Entries with low-confidence or unresolved class assignments, or labels spanning multiple classes, are held in a separate staging area for review. Medical terms rejected during validation against standard vocabularies are logged separately.

Concept grounding links exposure and outcome definitions to standardized medical concepts in the OMOP vocabularies. For diagnosis-based outcomes, the system retrieves candidates from the Condition domain, which contains concepts for diseases and other clinical conditions. An LLM selects concepts to include in or exclude from each outcome definition. Concept identifiers outside the retrieved candidate set are rejected.

During analysis, a concept resolver maps names recorded in the data back to standard concepts. Drug exposure is identified using the study-specific set of active ingredients. Outcomes are identified from records matching the inclusion concepts or their descendants. Descendant relationships are determined from the concept hierarchies in the OMOP vocabularies. Records matching exclusion concepts or their descendants are not counted toward that outcome. These constraints restrict selection to existing vocabulary entries and make exposure and outcome identification traceable. They are intended to reduce incorrect inclusion caused by matching free-text terms.

The system supports three operating modes. It can expand the knowledge base alone or expand it before protocol generation and analysis. Alternatively, it can analyze data using an existing knowledge base without literature-based expansion. In analysis-only mode, a frozen snapshot, or previously saved version of the knowledge base, can be loaded. This supports repeated evaluations with a consistent knowledge base.

### Evidence-grounded protocol generation and locking

The Case Generator uses design elements from target trial emulation to structure observational study protocols. This approach uses a hypothetical randomized trial to guide observational study design^5^. Each protocol specifies the study drug, comparator, target population, outcomes, and follow-up window. Exposure is defined at the drug-class or active-ingredient level. The system supports adverse-effect exploration, drug comparisons, and comparisons between different product strengths of the same drug. It also assesses whether drug–outcome associations differ across patient subgroups.

An active-comparator group receives an alternative treatment. A never-user group has no recorded exposure to the study drug. The choice depends on the research question and the clinical suitability of candidate comparator drugs. Comparator feasibility is assessed during cohort construction. Inclusion and exclusion criteria are tailored to the research question and draw on literature-derived candidates stored in the knowledge base. The protocol also specifies requirements for a new-user design, which focuses on patients initiating treatment. These include a washout period with no recorded use of the relevant drug before treatment initiation.

Outcome selection combines a core set with an exploratory extension. The core set includes primary outcomes specified in the research question. It also includes outcomes linked to existing drug-class evidence and serious outcomes from the safety panel. The Case Generator then identifies organ systems that the drug could plausibly affect based on its mechanism of action. It selects exploratory outcomes from the corresponding knowledge-base candidate pools. Selections outside these pools are excluded. Each outcome’s selection source is recorded in the protocol, and duplicates are merged. This combines question-driven analysis, prespecified safety surveillance, and mechanism-guided exploration to support proactive pharmacovigilance across multiple outcomes.

Covariate extraction uses a predefined set of patient characteristics informed by phenome-wide association studies (PheWAS). These studies examine associations across many health outcomes. The set includes demographics, healthcare use before study entry, comorbidity burden, and the length of medical history available before study entry. Comorbidity burden is summarized using an Elixhauser score, which combines pre-existing conditions using van Walraven weights^20^.

These covariates form a common pool for propensity score estimation. A propensity score estimates the probability of receiving the study treatment given recorded patient characteristics. Using a common pool supports consistent basic adjustment across outcomes and reduces discretion in choosing adjustment variables for individual questions. The covariates used and their availability are recorded during execution.

The protocol lock constrains analysis execution according to the generated study protocol. Before the Analyst begins patient-level analysis, the system saves the complete structured protocol. It also adds a timestamped record of key design fields to the audit log. These records support later checks of whether the analysis followed the protocol. The Case Generator is instructed to omit expected effect sizes and predictions about effect direction from the protocol. This is intended to reduce the influence of anticipated results on analytical decisions.

The locked protocol is made available to the analysis tools through a shared analysis context. Parameter inheritance and consistency checks take place when the relevant tools are called. When called, cohort tools inherit the protocol’s drug-class or ingredient-level exposure definition and its specified ingredient set. They also inherit protocol-specified inclusion and exclusion criteria, age limits, and washout requirements. The Analyst does not need to re-enter these settings.

The relevant tools also check the study drug class, comparator, outcome set, and follow-up window against the locked protocol. They reject conflicting calls, including attempts to add or remove protocol-specified outcomes. An error is returned to the Analyst so that it can correct the call. Main and subgroup analyses use the same protocol-specified follow-up window. These constraints govern key design decisions while retaining the Analyst’s discretion over execution details, including the sequence of tool calls, covariate selection for the propensity score model, and adjustment of the matching caliper.

When the comparator is infeasible, predefined feasibility rules within the tools govern fallback or termination. The comparator actually used and the reason for any fallback are recorded separately. The original protocol, analysis outputs, and checkpoint decisions are retained. Together, these records allow the executed analysis to be checked against the planned design.

### Tool-augmented analysis and checkpoint control

The Analyst is an autonomous tool-calling agent equipped with a library of pharmacovigilance analysis tools. It uses the locked protocol and outputs from preceding tools to select subsequent calls. Within the permitted scope, it can adjust analytical parameters or correct invalid calls. Human-verified deterministic tools perform patient-level numerical analyses. They return structured results, diagnostic information, and execution status. The Analyst coordinates the workflow within protocol constraints and the input requirements of each tool. New analytical methods can be incorporated by adding tools.

The cohort tools construct new-user cohorts according to the protocol’s exposure definitions and eligibility criteria. They support active-comparator and never-user designs, as well as comparisons of different product strengths of the same drug. Baseline covariate tools provide the predefined PheWAS-informed covariate set for propensity score estimation.

The matching tool performs 1:1 nearest-neighbor matching on the logit, or log-odds, scale of the propensity score. Each treated patient is paired with the closest available comparison patient. Matching is without replacement, so comparison patients are not reused. For patient-subgroup questions, initial matching is restricted to patients in the same subgroup, using each patient’s own subgroup label. Covariate balance is assessed using SMDs. The tools report patient losses during cohort construction and matching, together with any remaining covariate imbalance. These diagnostics help the Analyst assess feasibility and interpret the results.

Outcome ascertainment tools identify qualifying events during follow-up using standardized concept sets. Some outcome definitions require excluding prevalent cases, meaning patients with that outcome recorded at or before study entry. For these outcomes, the tools define an outcome-specific risk set, the patients eligible for that outcome analysis. Only complete original matched pairs in which both patients are eligible are retained. Covariate balance is reassessed within each risk set, without rematching.

Event ascertainment and censoring use the same follow-up boundaries. The cutoff is the earlier of the protocol-defined follow-up limit and the patient’s recorded observation-period end. Patients without a recorded event are censored at this cutoff, marking the end of their observation. The tools separately record outcomes that cannot be assessed, assessed outcomes with no observed events, and failed statistical estimates.

Association estimation tools use logistic regression to estimate ORs and Cox proportional hazards regression to estimate HRs, with CIs. Outcome models adjust for age and sex, while matching provides the primary control for confounding. When events are few or conventional fitting is unstable, logistic regression uses Firth penalized likelihood^21^. This method reduces small-sample estimation bias. Cox regression uses cluster-robust standard errors at the matched-pair level to account for dependence within pairs.

Multiple testing across outcomes is addressed separately for OR and HR analyses using the Benjamini–Hochberg procedure^22^. This procedure is used to control the expected proportion of false-positive findings among findings declared significant. Stratified analysis tools estimate associations within patient subgroups. Formal interaction tests assess whether these associations differ across subgroups. The tools also check model fits and record the status and reason whenever estimation or testing is not completed.

The checkpoint engine oversees the Analyst’s tool calls during execution. It receives results from tools for cohort construction, matching, balance assessment, and statistical analysis. Alongside their results, these tools return quality checks with measured values, status indicators, descriptions of issues, and recommended actions. Checks cover patient numbers in each study arm, the proportion retained after matching, and covariate balance. They also cover outcome event counts and whether statistical estimation succeeded. The tools assign quality statuses using predefined rules. The checkpoint engine uses these statuses to determine whether execution can continue.

In fully automated mode, the checkpoint engine allows steps that pass the checks to proceed. Warnings are recorded without stopping execution. Critical gate failures trigger an abort. For example, a tool can trigger an abort if a comparator cohort or matched sample fails predefined feasibility checks.

The tool wrapper links an analysis tool to the checkpoint engine. It records the abort decision in the shared analysis state. Subsequent analysis calls are then rejected. Termination therefore does not depend solely on the Analyst following a stop instruction. When continuation is permitted, tool diagnostics guide the Analyst’s subsequent calls and adjustments within the allowed scope. Skipped outcomes and estimation failures remain documented in the results and do not automatically terminate the entire study.

In human-in-the-loop mode, checkpoints present the current step’s metrics, issues, and recommendations to the researcher. The researcher decides whether to continue and can provide feedback. This feedback is returned to the Analyst with the tool output. It allows the researcher to guide subsequent execution while the analysis is in progress.

Each checkpoint record includes the step, status, decision, researcher feedback, and a timestamp. Together with the original protocol and analysis outputs, these records support review of key analytical decisions and the reasons for termination.

### Two-stage reporting and evidence comparison

MedCheckpoint uses two-stage reporting, with statistical results presented before comparison with existing evidence. In the first stage, the Report Writer is instructed to use the locked study protocol and structured analysis outputs. It writes results sections on the study population, covariate balance, and outcome analyses. Effect estimates, CIs, and result counts are taken from the analysis tool outputs. The agent adds each section to the draft using section writing tools. The writing instructions place literature retrieval and comparison after completion of these results sections.

In the second stage, the Report Writer queries the knowledge base for prior evidence and its sources. It compares the reported associations with prior findings in terms of direction and magnitude. The comparison considers the original study designs, effect measures, and certainty of the evidence. Evidence certainty describes confidence in the reliability of the prior findings. The agent discusses agreement, discrepancies, and possible biological mechanisms.

Interpretations supported by the literature are presented in a separate evidence comparison section. Citations are linked to the claims they support, and gaps in the evidence are explicitly identified. This ordering is intended to reduce anchoring, in which known effects unduly influence the selection and presentation of current results. Medical knowledge can then inform the interpretation of exploratory findings. Current findings and literature-based claims can be reviewed separately.

The Report Writer is instructed to report all outcome analysis results and their uncertainty. It is also instructed to describe the main data and design limitations and interpret findings as observational associations. Agreement with the literature can support plausibility but does not establish causality. When saving the report, the system directly appends the locked structured study protocol. This allows the reported design to be checked against the original protocol.

### Benchmark and comparison systems

We constructed a purpose-built pharmacovigilance benchmark of 54 natural language questions across three task categories (Supplementary Table 1). The benchmark included 21 questions on adverse drug reactions and off-target effects, and 14 comparisons between drugs. Another 19 questions examined how associations varied by dose or patient subgroup. Here, off-target questions explore effects beyond a drug’s intended therapeutic action. The questions were developed with reference to drug, population, and outcome records in the study dataset. They covered both open-ended exploration and analyses of outcomes specified in the questions.

The benchmark included a targeted challenge set of seven prespecified trap questions. These were deliberately designed around known data limitations that constrained analytical feasibility. Some involved cases in which a valid comparator cohort was difficult to construct. Others involved target outcomes without usable records in the data. These questions tested whether systems recognized analytical limitations and restricted their conclusions accordingly.

The first comparison system, Baseline A, is a general-purpose autonomous coding agent. It has access to the same EHR data as MedCheckpoint. It receives the research question, general pharmacoepidemiological guidance, and output requirements (Supplementary Note 1). It selects analytical methods and writes and executes its own code. This code performs cohort construction, confounding adjustment, outcome analysis, and report generation. Baseline A does not have access to MedCheckpoint’s knowledge base or specialized analysis tools. This comparison evaluates the analytical performance of a general-purpose agent working directly with the study data.

The second comparison system, Baseline B, uses the same execution framework as Baseline A but receives more detailed methodological guidance. This guidance is derived from the prompts for MedCheckpoint’s Case Generator, Analyst, and Report Writer (Supplementary Note 1). It covers target trial design, new-user cohorts, eligibility criteria, and confounding control. It also covers association estimation, stratified testing, and interpretation of results.

Baseline B does not have access to MedCheckpoint’s knowledge base, specialized tools, protocol lock, or checkpoint execution mechanisms. It still performs analyses using code written by the agent. Comparing MedCheckpoint with Baseline B assesses the full system’s overall benefit over methodological guidance supplied through prompts alone. Comparing Baseline B with Baseline A assesses the contribution of more detailed methodological guidance.

Claude Code (version 2.1.198) provides the execution framework for both baselines. Their backbone LLM is specified in the run configuration. All three systems use the same Parquet data containing concept names. The baselines also have read-only access to the original OMOP vocabularies. They cannot access MedCheckpoint’s curated knowledge base, concept sets, or source code. Reports and structured analysis results from each system are used for subsequent evaluation.

### Experimental setup and evaluation procedures

MedCheckpoint, Baseline A, and Baseline B all used MiniMax-M3 as the backbone LLM. Temperature, which controls randomness in LLM responses, was set to 0.1. Each system was run once per question in each of three independent batches. The natural language question served as the task input.

MedCheckpoint operated in fully automated mode using a prebuilt knowledge base snapshot. The knowledge base was not expanded during execution, and no human checkpoint decisions were made. Each baseline run took place in a separate container, an isolated software environment with read-only access to the study data. The baselines were prohibited from retrieving external information online. Their tool-call and execution logs were retained and audited for compliance. We summarized observed run times. Summaries covered all retained runs and question–batch combinations completed by all three systems (Supplementary Table 15).

Reports and structured analysis outputs from all systems underwent the same preprocessing before evaluation. A normalization program converted their different output formats into a common data structure. Original values and additional information were retained for verification. The preprocessing pipeline computed report-level metrics, including outcome counts, missing CIs, and covariate counts. It also classified each run’s completion status. Only runs classified as having completed the analysis were included in report-quality assessment.

For report-quality evaluation, system identity fields were removed from the structured results. Systems were assigned random anonymous labels within each question and batch. The label-to-system mappings were stored separately. Each set of evaluation materials contained the research question, report text, normalized results, and precomputed report metrics.

Kimi-K3 (16 July 2026 release), DeepSeek-V4-Flash (31 July 2026 release), and MiniMax-M3 (1 June 2026 release) served as LLM judges. Their base temperatures were 1.0, 0.0, and 0.0, respectively. For each evaluation task, all three judges received the same materials and scoring instructions (Supplementary Note 2). Their judgments were reported separately.

Pointwise scoring assessed each report individually. The five dimensions were Design Alignment, Cohort Validity, Statistical Rigor, Outcome Validity, and Interpretive Integrity. Each dimension was rated on a 1–5 scale. Judges were required to justify their scores using evidence from the report or analysis results. The scoring rubric instructed judges not to infer analytical correctness solely from agreement or disagreement with published effect directions. The rubric also penalized unsupported causal claims and interpretations of unmeasurable outcomes as evidence of no risk. The judges returned scores and explanations in a structured format. Before saving the results, the program checked required fields, permitted score values, and the presence of explanations.

Run-to-run stability was assessed using report quality scores from the three batches, separately for each judge. Only questions with valid scores in all three runs were included for each system. For each question and system, we averaged the five dimension scores within each run. We then calculated the sample SD of these averages across the three runs. These question-level SDs were averaged to summarize each system’s stability. We also calculated and averaged question-level SDs separately for each quality dimension. Lower SDs indicated more consistent report quality scores.

Pairwise comparisons assessed two reports at a time. They were restricted to question–batch combinations with valid evaluation materials from all three systems. Each judge evaluated every anonymous pair twice, with the presentation order reversed for the second evaluation.

Consistent (system 1) and Consistent (system 2) indicated that both judgments favored the corresponding system. Weak (system 1) and Weak (system 2) indicated one judgment favoring the corresponding system and one tie. Winner flipped by order indicated that the two judgments favored different systems. No preference indicated a tie in both orders. These six categories were counted and reported separately. In Fig. 3c, the two Weak categories, Winner flipped by order, and No preference were combined as No consistent preference.

We assessed interjudge agreement using pointwise scores for 442 completed reports. We calculated two-way mixed-effects, single-rating ICCs. ICC(A,1) measured absolute agreement, whereas ICC(C,1) allowed for systematic differences in judge severity^23^. For individual dimensions, Krippendorff’s ordinal α assessed three-judge agreement, and linearly weighted Cohen’s κ assessed judge pairs^24,25^. Gwet’s AC2 provided a prespecified complement when scores were concentrated. It used the same linear weights as κ but a different chance-agreement model^26^. Both κ and AC2 retained the full 1–5 scale. We also reported exact agreement and the proportion of reports with an interjudge score range ≤1.

Percentile 95% CIs were obtained from 5,000 question-level bootstrap samples. Each question’s reports and ratings were resampled together, with judges treated as fixed. Additional measures and sensitivity analyses are described in Supplementary Tables 6–9.

For the prespecified trap questions in this challenge set, judges received additional reference materials describing known data limitations (Supplementary Note 3). These explained whether the intended comparison groups could be constructed and which outcomes were not expected to be reliably identified. They also provided the rationale for these expectations.

Judges evaluated reports or abort records against these materials using a common rubric. Each assessment was saved in a structured format containing the judgment, its justification, and the type of handling error, if any. Justifications had to cite specific evidence from the report or abort record.

We retrospectively audited analysis completion and termination across systems. Questions from the data-limitation tests were excluded. A question was selected if at least one system had a run initially classified as not completed during output preprocessing. For every selected question, we included all three runs from each system. The audit comprised 14 questions and 42 runs per system (Supplementary Table 11).

We reviewed original reports, analysis outputs, and execution logs, checking source data when needed. LLMs assisted with organizing the evidence, and human review determined the final judgments. Execution outcomes were classified separately from report quality. The three categories were Completed with available data, Properly stopped (stopped at cohort or matching checks), and Fabricated answer without analyzing.

Within this audit sample, we reused the completed reports’ existing pointwise scores to compare report quality. We first summarized each system’s completed reports. We then restricted the comparison to question–batch combinations completed by all three systems. Each report’s overall score was the mean of its five dimension scores. System means were calculated with equal weight per report, separately for each judge.

## Data availability

The data used in this study are publicly available through the Registry of Open Data on AWS (https://registry.opendata.aws/synthea-omop/). We used the largest available dataset, located in the synthea23m/ directory.

## Code availability

The code used in the article is available upon request to R.L.

## Acknowledgements

R.L. was supported by Hong Kong Research Grants Council GRF (17212626), Hong Kong Innovation and Technology Commission MHKTCFS (GHP/547/25GD), China NSFC Excellent Young Scientist Fund (32622025), and the URC fund at HKU.

## Author contributions

Y.Z., and R.L. conceived the study. R.Z. developed the methodology and performed the computational analyses. C.S.L.C., D.Y., Z.W., and I.C.K.W. analyzed the results. R.Z. and Y.Z. drafted the manuscript. Y.Z. and R.L. supervised the project. All authors reviewed, revised, and contributed to the final manuscript.

## Competing interests

I.C.K.W. received research grants outside of submitted work from European Commission, Research Grants Council of the Hong Kong, Health and Medical Research Fund of the Government of the Hong Kong and Amgen; consulting fees from IQVIA and World Health Organization; served as a member of Pharmacy and Poisons Board of Hong Kong; was the founder and director of Therakind Limited. He is the non-executive director of Jacobson Pharma Corp. Ltd. in Hong Kong; and director of Advance Data Analytics for Medical Science Limited (HK), Healthcare Innovation Technology Service (HITS) limited and OCUS Innovation Limited (HK, Ireland and UK). He received honorarium from Pfizer and Takeda as a speaker.

## Notes

### Competing Interest Statement

The authors have declared no competing interest.

### Summary of Updates

Author list updated; wordings corrected

